# The consistent burden in published estimates of delirium occurrence in medical inpatients over four decades: a systematic review and meta-analysis study

**DOI:** 10.1101/19005165

**Authors:** Kate Gibb, Anna Seeley, Terry Quinn, Najma Siddiqi, Susan Shenkin, Kenneth Rockwood, Daniel Davis

## Abstract

**Introduction:** Delirium is associated with a wide range of adverse patient safety outcomes. We sought to identify if trends in healthcare complexity were associated with changes in reported delirium in adult medical patients in the general hospital over the last four decades.

**Methods:** We used identical criteria to a previous systematic review, including studies using DSM and ICD-10 criteria for delirium diagnosis. Random effects meta-analysis pooled estimates across studies, meta-regression estimated temporal changes, funnel plots assessed publication bias.

**Results:** Overall delirium occurrence was 23% (95% CI 19%-26%) (33 studies). There was no change between 1980-2019, nor was case-mix (average age of sample, proportion with dementia) different. There was evidence of increasing publication bias over time.

**Discussion:** The incidence and prevalence of delirium in hospitals appears to be stable, though publication bias may mask true changes. Nonetheless, delirium remains a challenging and urgent priority for clinical diagnosis and care pathways.

## Introduction

Delirium – a major complication of dementia – is characterised by disturbance of consciousness and inattention triggered by an acute event (e.g. medical illness, surgery) [1]. It is substantially underdiagnosed in clinical practice, partly driven by its fluctuating nature and the diversity of clinical manifestations. It is associated with a wide range of adverse outcomes, particularly those relevant to patient safety. These include: mortality, falls, increased length of stay, and risk of institutionalisation [2, 3]. In longitudinal studies, dementia is the biggest risk factor for delirium, and reciprocally, delirium is linked with worsening cognitive decline and incident dementia [4, 5].

That delirium was a substantial burden among hospitalised older adults was established in a 2006 systematic review, describing delirium prevalence as ranging from 10 to 31% across 42 studies since 1980 (when delirium was first formally defined in DSM-III) [6]. Subsequently, a number of initiatives confirmed the need for better delirium prevention and management [7]. Increased focus on delirium coincided with secular changes in the average patient age, background hospital prevalence of dementia and higher care complexity that came with more frailty in patients admitted to hospital [8, 9] with a consequent need to develop appropriate services. These underlying trends would be expected to lead to increases in delirium presentations, though this has never been directly investigated. Contemporary estimates of delirium epidemiology are needed, with implications for identifying training needs, clinical practice and public health policy [10]. In view of this, we set out to update the original systematic review in order to describe any change in the prevalence or incidence of delirium in the context of healthcare developments over the last four decades.

## Methods

### Eligibility criteria

We used identical criteria to the previous review [6], in line with PRISMA guidance [11]. We considered prospective cohort and cross-sectional studies describing delirium in adults (aged 18 or older) who were acute, unscheduled admissions (including stroke and oncology patients) in any country and in any language. We did not include randomised controlled trials if we were unable to estimate cases in an unselected denominator. We excluded studies in terminally ill patients and those solely in patients referred to liaison psychiatry services.

Studies in purely surgical cohorts, psychiatric units, emergency departments, coronary and intensive care units were excluded; studies in mixed populations were included if they separately reported information on internal medicine inpatients. Settings outside acute hospitals were excluded including post-care units, rehabilitation units, hospices and specialist palliative care units, and community hospitals. Reports on delirium specific to a clinical setting were excluded: e.g. delirium tremens, emergence delirium, post-electroconvulsive therapy, post-head injury. We only included peer-reviewed publications (i.e. we excluded abstracts and grey literature).

### Outcome measures

We included only studies which diagnosed delirium according to the Diagnostic and Statistical Manual of Mental Disorders (DSM) or the International Statistical Classification of Diseases (ICD). To be included, ascertainment needed to have been performed by a person trained to apply the relevant reference standard (e.g. geriatrician, psychiatrist, nurse specialist, researcher); studies relying on routine clinical ascertainment were excluded. Studies where participants were pre-screened with a non-diagnostic tool prior to applying DSM or ICD to those screening positive for delirium were also excluded unless a sample of screen-negatives were also assessed.

Using an established operationalised reference standard is essential to investigate change over time, though different iterations of these classifications are inevitably also subject to temporal trends. Of the 42 cohorts included in the original review, we carried forward 15 studies that met this eligibility criterion [12-26].

In describing the epidemiology of delirium in hospitals, *prevalence* conventionally refers to delirium ascertained on admission, *incidence* refers to delirium developing at some point over the inpatient admission. Where these have been difficult to distinguish – due to delirium fluctuations and/or different frequencies of observation – the more neutral term *occurrence* has usually been used. We considered studies which assessed the prevalence, incidence or occurrence of delirium.

### Search strategy

Updating the original review, we searched from one year prior to the previous end date (July 2004) to 31^st^ May 2019. We searched the same electronic databases: Medline, EMBASE, PsycINFO, CINAHL Plus and the Cochrane Database of Systematic Reviews, using the following search terms; Delirium [Title] AND (epidemiology OR prevalence OR incidence OR occurrence) [Title/Abstract]. We confirmed the sensitivity of the search by ensuring all studies from the previous review were captured.

### Data collection and study selection

Covidence (www.covidence.org, Veritas Health Innovation Ltd.) was used to manage the abstract and full text screening, assessment of risk of bias and data extraction. Titles and abstracts were independently reviewed by two reviewers (KG, AS) to determine eligibility for inclusion. Conflicts were resolved by discussion and consensus. Data extraction for primary outcome and key variables was also performed by two reviewers (KG, AS or DD) using a pro forma.

### Assessment of quality and biases

There is no consensus on the best tool for assessing risk of bias in descriptive epidemiology. The previous review used adapted criteria developed by the original authors.[27] We extended this previous approach by also accounting for items referred to in the Standards of Reporting of Neurological Disorders (STROND) criteria [28, 29]. Ultimately, we considered five domains: (i) patient setting, e.g. general medical versus stroke patients; (ii) sample selection, e.g. randomised or convenience approach; (iii) sample criteria, e.g. exclusions based on capacity to consent or language; (iv) use of a defined reference standard; (v) expertise of assessor applying reference standard. Each criterion was independently graded as low, medium or high risk of bias and we visualised this using the *robvis* package [30]. We described certainty of our findings using an approach based on the GRADE framework, where we assessed risk of bias; consistency of results (based on heterogeneity); directness (applicability of included studies to research question); precision (based on confidence intervals of summary estimate) and publication bias (based on funnel plot) [31].

### Statistical analyses

We extracted summary statistics for prevalence, incidence and occurrence, along with their standard errors. We anticipated methodological heterogeneity across cohorts, so accounted for this by calculating pooled estimates using DerSimonian-Laird random effects models [32]. Statistical heterogeneity was assessed with the I^2^ statistic. Meta-regression was used to estimate change over time and we used linear regression to examine if studies varied in average age or dementia prevalence in the samples, by year of publication. To assess publication bias, we plotted the estimated proportion of delirium occurrence against the standard error of that estimate, with Egger regression quantifying the degree of asymmetry. Stata 14.1 (StataCorp, Texas) was used for all analyses.

## Results

The search identified 4137 citations of potential relevance. After removing duplicates, we screened 3093 titles and abstracts, and assessed 189 for full text review for eligibility (Figure 1). Full text screening excluded 171 studies; 50 were conference abstracts, 52 used methods other than DSM or ICD to diagnose delirium, 14 were studies of patient population not of interest, e.g. surgical, intensive care patients. All reasons for exclusion are detailed in Figure 1. We included 18 studies [33-47], adding to 15 studies from the original review.

**Figure 1:**
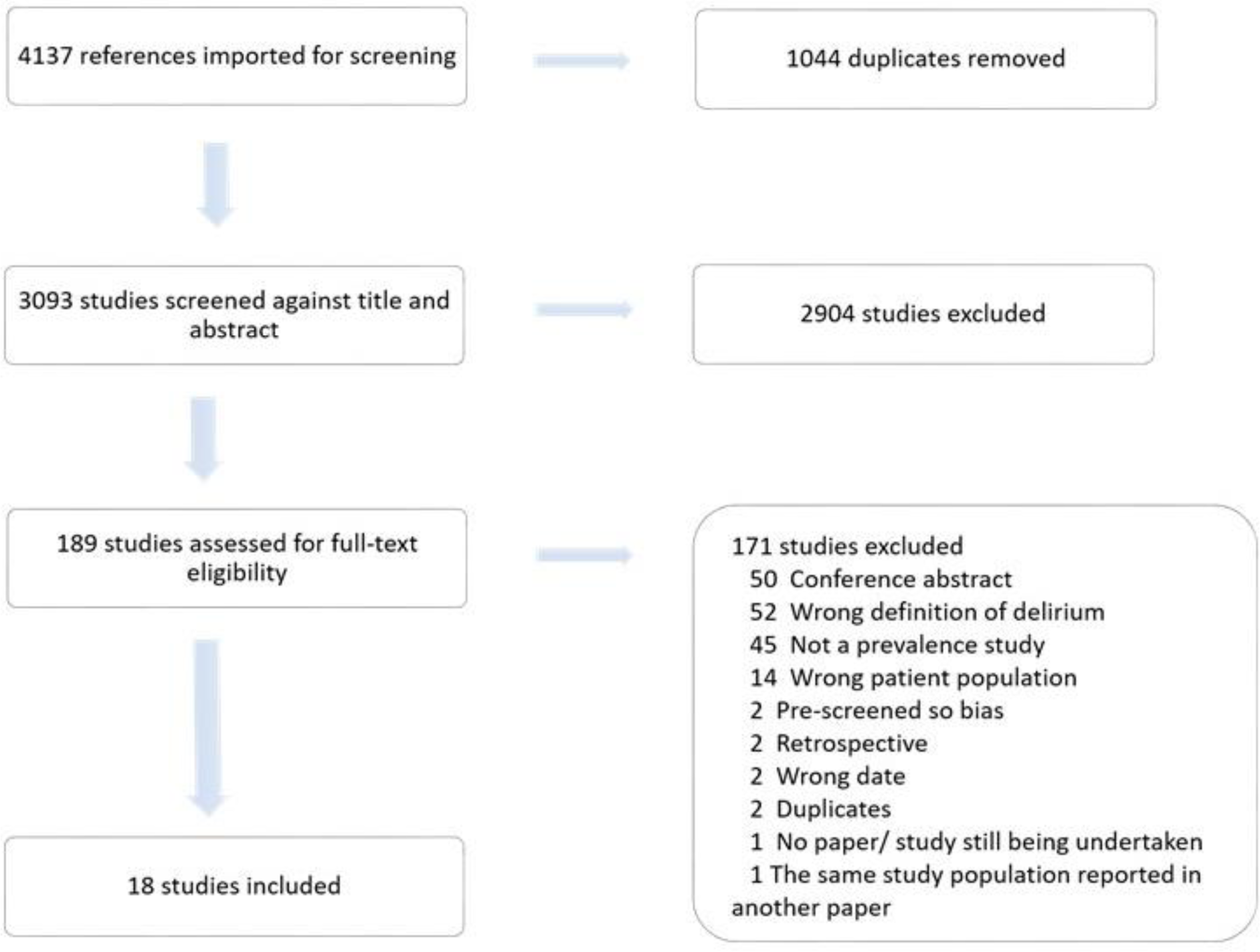
PRISMA flowchart of search results and study retrieval.

### Study characteristics

All studies were carried out in acute medical or geriatric medicine units, and all were prospective cohort studies, except one cross-sectional study (Table 1). Most were conducted in Western European populations, though single studies from China, Turkey and Thailand were included. Studies ranged in size from n=60 to n=1327, and varied in age (range of average sample age from 66 to 87 years) and prevalence of co-morbid dementia (range 8% to 100%). Delirium was diagnosed using DSM-IV or DSM-5 in sixteen studies, and two used ICD-10, adding to the six using DSM-III, six using DSM-III-R and three using DSM-IV from the original review. Some studies reported estimates based on more than one criterion, therefore 35 occurrence estimates are included in Figure 2.

**Table 1.** Characteristics of included updated studies.

| Study | Country | Sample | Exclusion criteria | N | Mean age (yrs) | Dementia prevalence | Reference standard |
| --- | --- | --- | --- | --- | --- | --- | --- |
| Adamis, 2015 | Ireland | ≥ 70 years; all acute medical admissions. | Hospitalised for > 48 hours; readmitted to unit; studied on previous admission; severe aphasia; intubated; sensory problems; non-English speaking | 200 | 81.1 | 63% | DSM-IV, DSM-5 |
| Bellelli, 2018 | Italy | ≥ 70 years, consecutive admissions (multiple hospitals) | No proxy available for consent | 588 | 80.9 | 12% | DSM-5 |
| Bonetti, 2012 | Italy | > 64 years; admissions to geriatric units | Nil | 578 | 82 | NR | DSM-IV |
| Chan, 2016 | China | ≥ 18 years; admissions to the respiratory wards for acute respiratory failure with non-invasive positive pressure ventilation. | Persistent coma; those who lacked mental capacity to provide consent and guardian not available; unavailable in first 48 hours of admission (died or discharged) | 153 | 74.2 | 7.8% | DSM-IV |
| Grandahl, 2016 | Denmark | ≥ 18 years; admission to oncologic ward; histologically verified cancer diagnosis | Non-Danish speaking patients; readmitted to unit; studied on a previous admission. | 81 | 68.5 | NR | ICD-10 |
| Holttä, 2015 | Finland | ≥ 70 years; admissions to acute geriatrics wards | Coma | 255 | 86.6 | 100% | DSM-IV |
| Jackson, 2016 | UK | ≥ 70 years; admissions to acute medicine | Unable to communicate because of severe sensory impairment; unable to speak English; at risk of imminent death. | 1327 | 84.4 | 36% | DSM-IV |
| Kozak, 2016 | Turkey | ≥ 18 years; clinical presentation of acute ischaemic stroke | Admission to hospital after first 24 hours; a diagnosis of TIA, cerebral haemorrhage; reduced GCS, severe aphasia or dysphasia; history of brain tumour, myocardial infarction, infection, autoimmune and immunosuppression, recent trauma or surgery; renal dysfunction and symptomatic peripheral arterial disease; GI or rheumatic inflammatory disease, metabolic syndrome; recent antidepressant use. | 60 | 66.2 | NR | DSM-IV |
| Laurila, 2004 | Finland | ≥ 70 years | Coma | 219 | ≥85=59% | 40% | DSM-IV |
| Paci, 2008 | Italy | Stroke; admissions to the Stroke Unit during the first 5 days of hospitalisation. | Nil | 150 | 67.5 | NR | DSM-IV |
| Pendlebury, 2015 | UK | Admissions to acute medical unit | Nil | 503 | 72 (median) | 10% | DSM-IV |
| Pitkala, 2004 | Finland | ≥ 70 years | Coma | 230 | ≥85=62% | 61% | DSM-IV |
| Praditsuwan, 2012 | Thailand | ≥ 70 years; admissions to general medical wards | Endotracheal intubation at admission; aphasia; uncooperative; coma | 225 | 78 | 42% | DSM-IV |
| Sheung, 2006 | Australia | ≥ 65 years; admissions with acute stroke | TIAs; subarachnoid haemorrhage; history of severe head trauma or neurosurgery before stroke; stroke due to tumour or cerebral venous sinus thrombosis | 156 | 79.2 | 7.7% | DSM-IV |
| Thomas, 2012 | Germany | ≥ 80 years; admissions to geriatric unit | Global aphasia; terminal condition | 79 | 84.1 | 75% | DSM-IV, ICD-10 |
| Travers, 2012 | Australia | ≥ 70 years; admissions to general medical and surgical wards; expected hospitalisation > 48 hours | Transferred to a study ward from another hospital or ward and admitted for > 48 hours previously; immunocompromised; imminent death | 294 | 80.4 | 26% | DSM-IV |
| Uchida, 2015 | Japan | ≥ 65 years; incurable lung or GI cancer; planned admission of ≥ 2 weeks; Performance Status of 2 or worse | Physically too ill to complete the survey; non-Japanese speaking | 61 | 72 | NR | DSM-IV |
| Yam, 2018 | China | ≥ 65 years; admissions to general medical wards | Direct admissions to the intensive care unit, coronary care unit and acute stroke unit; coma, persistent vegetative state; severe aphasia; clinically unstable; deemed too unwell | 575 | 80.8 | NR | DSM-5 |

NR: not reported. Note some sample overlap is possible between Pitkala 2004 and Laurila 2004.

**Figure 2.**
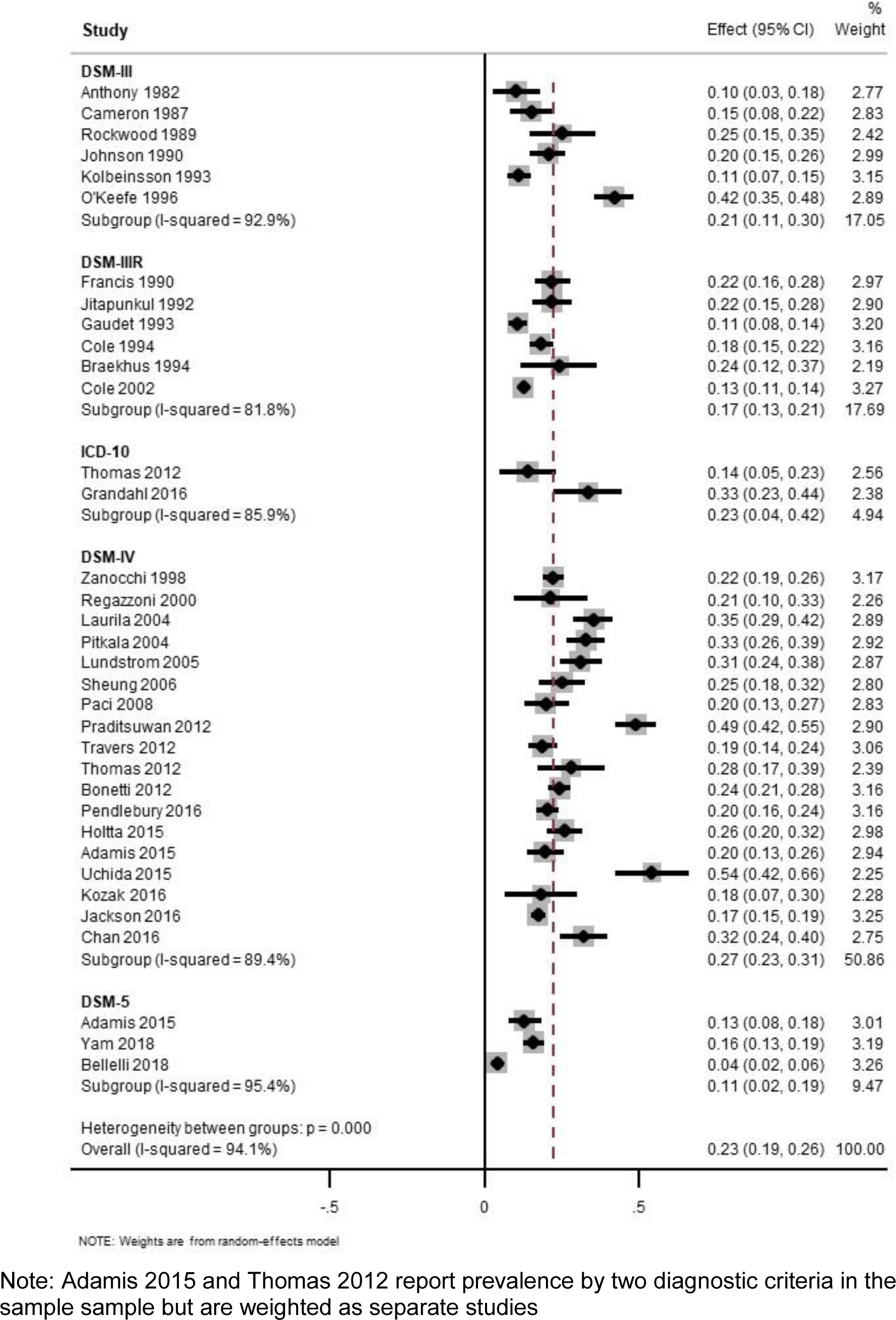
Meta-analysis of included studies (with studies from original review), stratified by diagnostic criteria and ordered by publication date. Note: Adamis 2015 and Thomas 2012 report prevalence by two diagnostic criteria in the sample sample but are weighted as separate studies

### Study quality

Sources of risk of bias were assessed in all studies (including from the original review) according to the domains detailed in Figure 3. Studies scored “low risk” or “some concerns” in all domains, with 27 of 33 studies considered to be low risk overall. Most studies were rated “some concerns” for source population because the sample was from a single centre (Domain 2, Figure 3). Other studies had potential sources of bias through excluding people with severe aphasia, inability to communicate due to severe sensory problems, those lacking capacity to consent (or no provisions for proxy consent), terminally ill, or in coma (Domain 3, six studies).

**Figure 3.**
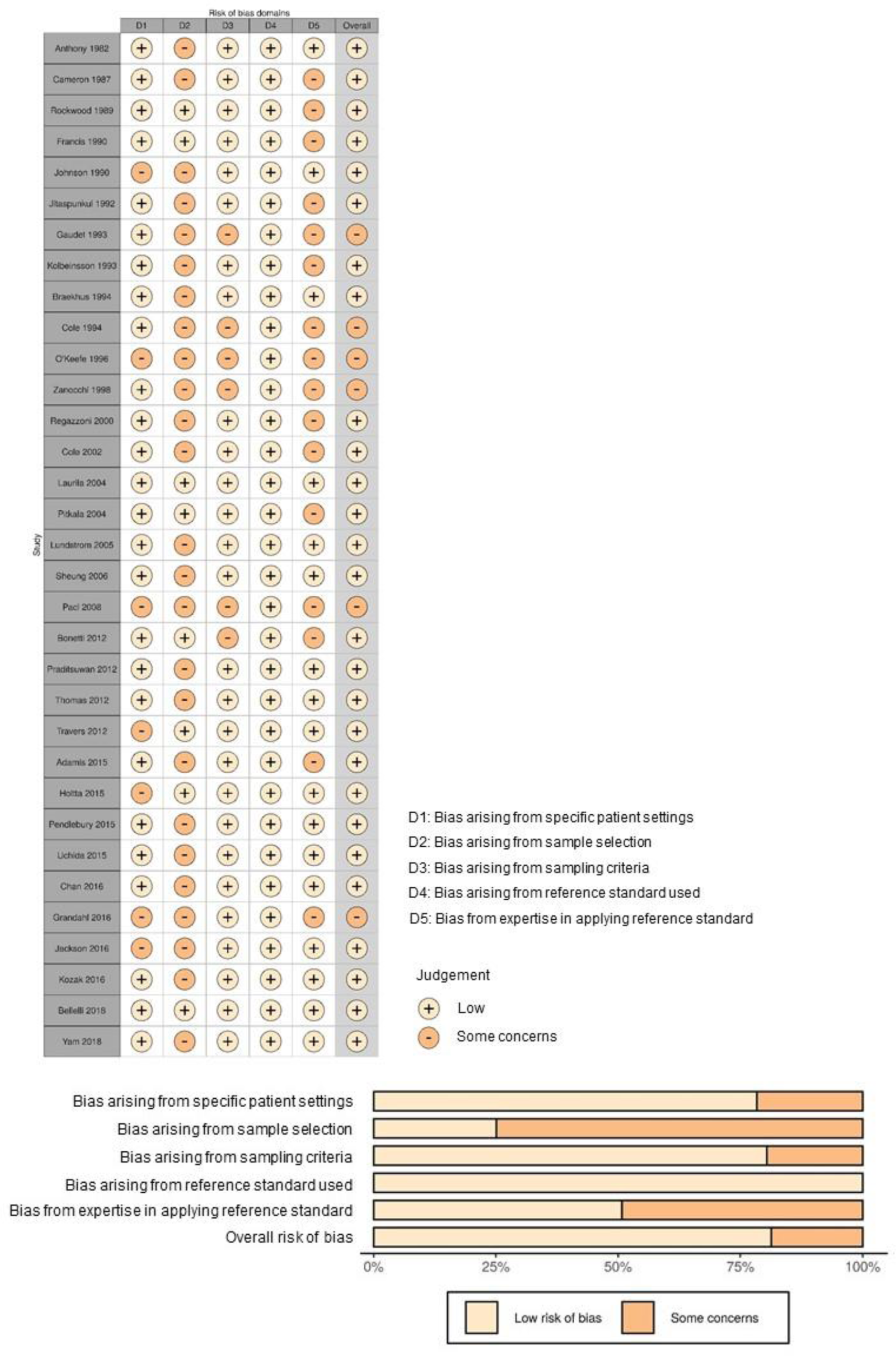
Risk of bias assessments.

### Delirium prevalence, incidence and occurrence

Pooled prevalence was estimated as 15% (95% CI 14% to 16%, 25 studies). Cumulative incidence of new delirium was 9% (95% CI 7 to 10%, 14 studies) over the observed period, which was up to two weeks in duration. Figure 2 shows estimates of total delirium occurrence of 23% (95% CI 19% to 26%), stratified by reference standard. Differences in occurrence estimates were evident according to diagnostic criteria, with DSM-IV and DSM-5 showing higher and lower estimates respectively.

Figures 4a-c indicate the prevalence, incidence and occurrence over time (1980 to 2018). Meta-regression models did not demonstrate any statistically significant temporal changes (prevalence: increasing by 0.2%/year, 95% CI -0.2% to 0.6%/year, p=0.38; incidence: - 0.1%/year, 95%CI -0.4% to 0.4%/year, p=0.95; occurrence: 0.2%/year, 95%CI -0.2% to 0.5%/year, p=0.35).

**Figure 4a-c.**
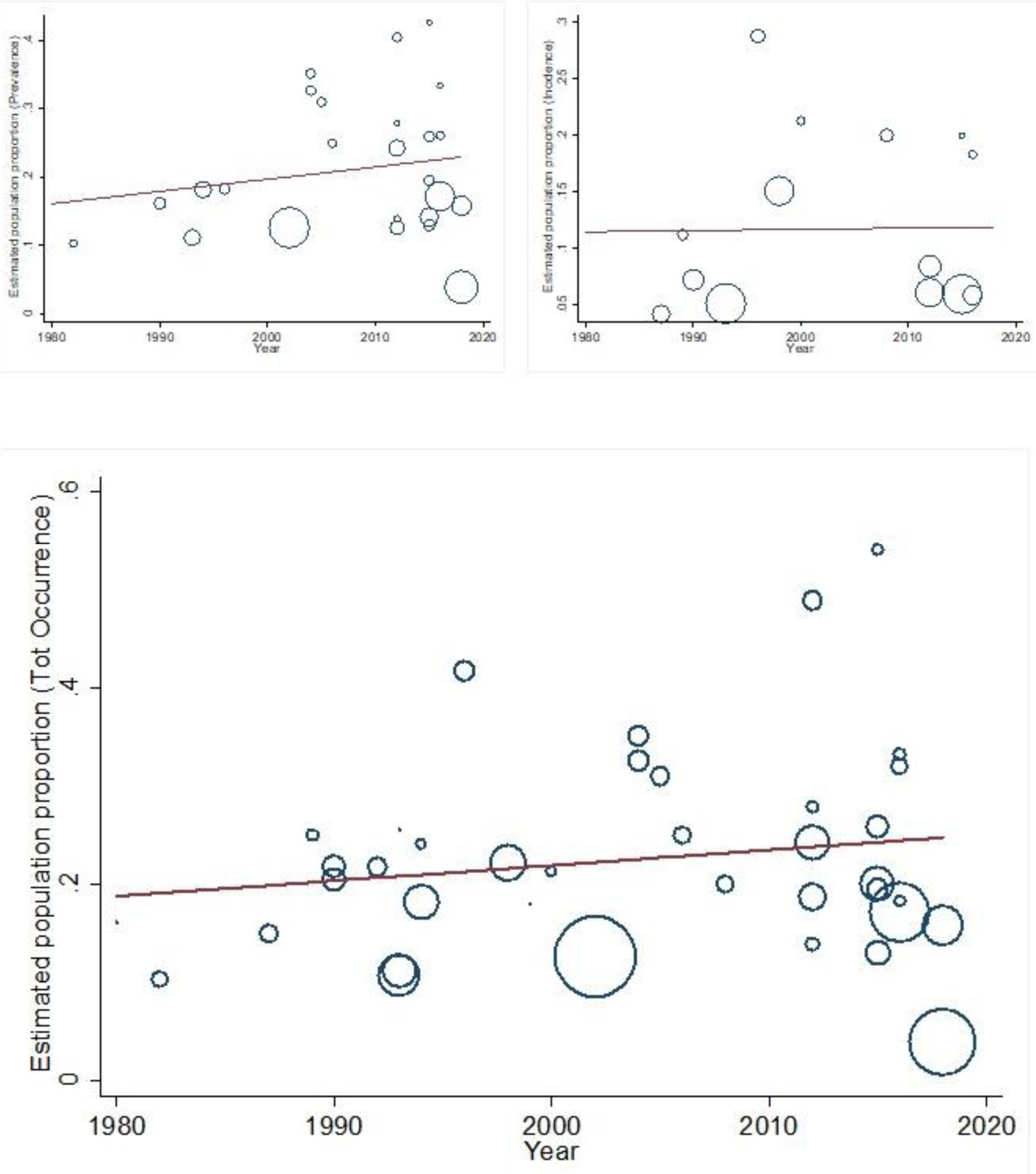
Temporal trends in delirium prevalence (top left), incidence (top right) and occurrence (bottom).

Over time, there were no differences in the average age of the samples in included studies (mean age across studies 80.0 years, change over time -0.28/year, 95% CI -0.79 to 0.24, p=0.28). Where studies indicated the prevalence of comorbid dementia in the sample (n=19), these also did not show any secular changes over the study period (mean prevalence of dementia 40%, change over time 0.11%/year, 95% CI -0.02% to 0.23%, p=0.10).

### Publication bias

Publication bias was suggested from asymmetry in forest plots (Egger coefficient 5.10, p<0.01, Figure 5). However, this was not apparent in earlier studies and each successive decade was associated with more funnel plot asymmetry (1980-1989 coefficient 4.24, p=0.32; 1990-1999 coefficient 4.22, p=0.09; 2000-2009 coefficient 5.08, p=0.02; 2010-2019 coefficient 5.99, p=0.01).

**Figure 5.**
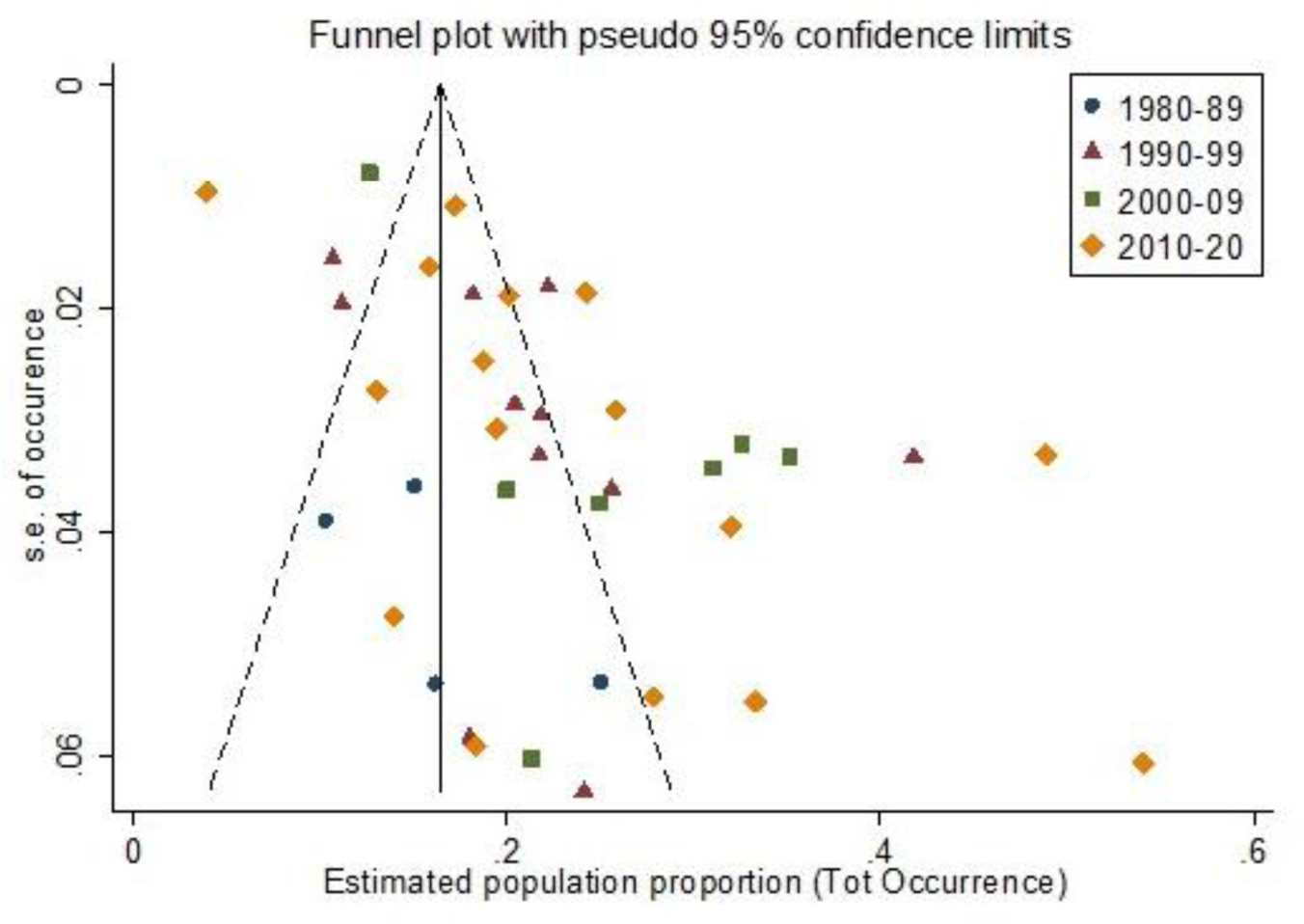
Funnel plot showing occurrence of delirium in relation to standard error of the estimate, by decade.

## Discussion

In the last four decades, the published prevalence and incidence of delirium in acute medical adult inpatients has remained broadly stable at 23% (95% CI 19%-26%). We quantified this from studies using consistent methods in comparable populations. There were no major differences in case mix (average age, dementia prevalence) across time. However, there was evidence for increasing publication bias, suggesting that estimates confirming a higher apparent burden of delirium are more likely to be published; these samples may not be representative of clinical patients in routine care. Taken together, delirium remains a substantial problem in acute hospitals, though quantifying this in relation to increased healthcare complexity alongside increased prioritisation of delirium in clinical practice is not straightforward.

Several limitations to our findings require further comment. To be consistent with the original review, we only considered studies on acute medical and geriatric medicine inpatients. This limits generalisability to other settings. We could not account for illness severity nor were direct measures of frailty available. While it is clear that most delirium risk is conferred by age and baseline dementia status, it is possible that more nuanced measures may have captured changes in case mix more accurately. We expected to see variation in case mix across time; at least for average age and underlying dementia prevalence, this did not appear to change. Publication bias might reflect this to an extent – geriatric medicine wards in the earlier studies may have been comparatively less common and more likely to lead to a report on delirium prevalence. Publication bias certainly appears to account for some of the funnel plot asymmetry, where more recent studies report higher occurrence than might be expected by virtue of their size. If as a consequence, these are less representative of clinical patient populations, then prevalence and incidence of delirium may be being overestimated in our included studies. Other aspects to the risk of bias assessments indicated that our findings were not subject to much variation due to training of the diagnostic rater, or limited by much selection bias because of inappropriate exclusions.

To highlight the overall clinical implications, no net change in the reported epidemiology confirms delirium as a major public health concern affecting approximately 1 in 4 adults admitted to an acute general hospital. In particular, rates of incident delirium remain high, suggesting that front-door preventative measures have not made substantial impact.

However, there is also the possibility that diverging trends underlie our findings. On the one hand, increasing complexity of healthcare and frailty among acute admissions may lead to more delirium. In contrast, delirium has attracted much more prominence in recent years with increased emphasis on multicomponent prevention [48], representation in clinical care pathways and guidelines [49] and recognition of its potential role in dementia prevention [10]. There is some suggestion this may have been effective in the context of acute stroke services [50]. However, if the publication bias leads to inflated estimates of delirium occurrence in more general settings, then the effectiveness of these trends may be being masked. Nonetheless, it is clear the extent of delirium remains considerable. There can be no complacency around prioritising the entire delirium care pathway, from risk recognition, diagnosis, prevention and management.

In this updated systematic review and meta-analysis, we found the epidemiology of delirium among hospitalised patients has not changed substantially between 1980 and 2018. At least in estimates from the published literature, case mix also appears not to have changed much. With this burden of delirium in hospitals, contemporary priorities around disseminating delirium knowledge, increasing the proportion diagnosed and implementing care pathways remain as challenging yet urgent as ever.

## Data Availability

Data used in this manuscript are publicly available.

## Acknowledgements

Kate Gibb and Anna Seeley are supported through UCLH CEO Clinical Research Training Fellowships. Daniel Davis is supported through a Wellcome Trust Intermediate Clinical Fellowship (WT107467).

